# Detection of SARS-CoV-2 nucleocapsid antigen from serum can aid in timing of COVID-19 infection

**DOI:** 10.1101/2021.01.08.20248771

**Authors:** MJ Ahava, S Kurkela, S Kuivanen, M Lappalainen, H Jarva, AJ Jääskeläinen

**Author notes:** Corresponding author: Maarit J Ahava, Helsinki University Hospital, HUSLAB, Virology and Immunology, P.O.B. 720 (Topeliuksenkatu 32), FI-00029 HUS, Finland.

## Abstract

SARS-CoV-2 RNA can be detected in respiratory samples for weeks or even months after onset of COVID-19 disease. Therefore, one of the diagnostic challenges of PCR positive cases is differentiating between acute COVID-19 disease and convalescent phase. Recently, the presence of SARS-CoV-2 nucleocapsid antigen in serum samples of COVID-19 patients was published [Le Hingrat et al. Detection of SARS-CoV-2 N-antigen in blood during acute COVID-19 provides a sensitive new marker and new testing alternatives, *Clinical Microbiology and Infection*, 2020].

Our study aimed to characterize the analytical specificity and sensitivity of an enzyme-linked immunosorbent assay (Salocor SARS-CoV-2 Antigen Quantitative Assay Kit^©^ (Salofa Ltd, Salo, Finland)) for the detection of SARS-CoV-2 antigen in serum, and to characterize the kinetics of antigenemia. The evaluation material included a negative serum panel of 155 samples, and 126 serum samples from patients with PCR-confirmed COVID-19.

The specificity of the Salocor SARS-CoV-2 serum N antigen test was 98.0%. In comparison with simultaneous positive PCR from upper respiratory tract (URT) specimens, the test sensitivity was 91.7%. In a serum panel in which the earliest serum sample was collected two days before the collection of positive URT specimen, and the latest 48 days after (median 1 day post URT sample collection), the serum N antigen test sensitivity was 94% within 14 days post onset of symptoms. The antigenemia resolved approximately two weeks after the onset of disease and diagnostic PCR.

The combination of simultaneous SARS-CoV-2 antigen and antibody testing appeared to provide useful information for the timing of COVID-19. Our results suggest that SARS-CoV-2 N-antigenemia may be used as a diagnostic marker in acute COVID-19.

## Introduction

SARS-CoV-2 laboratory diagnostics relies primarily on molecular diagnostic techniques [1-3]. More recently, a variety of tests for SARS-CoV-2 antigen detection from upper respiratory tract (URT) specimens have established a complementary role [4]. While less sensitive, they benefit from being rapid, cheap, and performable outside centralized laboratory facilities.

Recent articles and preprints demonstrate SARS-CoV-2 nucleocapsid (N) antigenemia in COVID-19 patients [5,6]. While posing new questions on the pathophysiology of acute COVID-19, this may offer a novel diagnostic approach. Only limited information is available on the performance of serum antigen tests as a diagnostic method for SARS-CoV-2.

The aim of this study was to characterize the analytical specificity and sensitivity of an enzyme-linked immunosorbent assay (ELISA) for the detection of SARS-CoV-2 antigen in serum, namely Salocor SARS-CoV-2 Antigen Quantitative Assay Kit^©^ (Salofa Ltd, Salo, Finland; later Salocor N-antigen EIA) and to characterize the kinetics of antigenemia.

## Materials and methods

Serum samples were originally sent for diagnostic purposes to the Department of Virology and Immunology, Helsinki University Hospital Laboratory HUSLAB, Finland. Research permit HUS/157/2020-44 (Helsinki University Hospital, Finland) was obtained from the local review board.

The negative panel (N=155; 148 cases) consisted of 144 serum samples collected in 2019, and 11 samples (positive for Aspergillus antigen) in 2020. Of the 155 specimens, 37 were sent for respiratory virus antibody testing; 32 samples were positive for anti-nuclear antibodies; 16 for phospholipase-A2-receptor antibodies; 8 for antineutrophil cytoplasmic (C-ANCA, P-ANCA and parallel C- and P-ANCA), and 4 for glomerular basement membrane antibodies. Two samples were from patients with Human coronavirus (HCoV) OC43 diagnosis (by PCR) and five with primary Epstein-Barr virus (EBV) infection. We also included serum specimens with a positive microbial antigen test as follows: 22 with Dengue virus NS1 antigen, 17 with hepatitis B virus surface antigen, 11 with Aspergillus antigen, and one with HIV p24 antigen. The median age of the negative panel cases was 53 years (range 2-89); 45% were males (66/148).

There were two separate serum specimen panels from PCR confirmed COVID-19 patients (panel A and panel B), who were previously SARS-CoV-2 PCR positive from a URT specimen. They were also tested for SARS-CoV-2 IgG by both Abbott SARS-CoV-2 IgG (N antigen) and Euroimmun SARS-CoV-2 IgG (S1 antigen) according to manufacturer’s instructions.

Panel A comprised 70 serum samples from 62 cases (median age 54 years, range 24-86 years; 28/62 (45%) males). The earliest serum sample was collected 2 days before the collection of the positive URT specimen, and the latest 48 days after (median 1 day post URT sample collection). The samples were previously tested with SARS-CoV-2 microneutralization (MNT) [7]. The date of onset of symptoms was available for 55/62 cases. Adjusted p-values for comparison of antigen and MNT result combinations were determined using Kruskal-Wallis test (GraphPad Prism 8.0.1).

Panel B comprised 56 serum samples from 27 cases (median age 50 years, range 27-65 years; 4/27 males); 2-4 consecutive serum samples from each. At least one serum from all 27 cases was SARS-CoV-2 IgG positive in both Abbott and Euroimmun tests. The earliest serum sample was collected 5 days before the collection of the positive URT specimen, and the latest 207 days after (median 101 days post URT specimen collection). The date of onset of symptoms and MNT result were not available for this panel.

The PCR tests were carried out with one of the following methods: a laboratory-developed test based on Corman et al.; cobas® SARS-CoV-2 test kit on the cobas® 6800 system (Roche Diagnostics, Basel, Switzerland); and the Amplidiag® COVID-19 test on the Amplidiag® Easy platform (Mobidiag, Espoo, Finland). The performance of these tests in our laboratory is reported elsewhere [8].

The here-evaluated Salocor N-antigen ELISA (Salofa) is based on a double antibody sandwich ELISA test. The assay protocol is described in the Supplement.

The 95% Clopper-Pearson confidence intervals were calculated for sensitivity and specificity (IBM SPSS statistical program package, version 25).

## Results

The specificity of the Salocor N antigen ELISA was 98.0% (145/148) (Clopper-Pearson 95% confidence interval 94.2-99.6%) as determined by the negative panel. The three positive samples were collected in 2019. One was from a patient with EBV primary infection (5 pg/ml). Two were originally sent for respiratory virus antibody screening; one with reported myocarditis (84.5 pg/ml) and one without any reported clinical outcome (4 pg/ml).

There was a simultaneous serum and PCR positive URT specimen available in 24 cases: N antigen was positive in 22/24, rendering 91.7% sensitivity (Clopper-Pearson 95% confidence interval 73.0-99.0%). The negative specimens were retrieved at one day and at 16 days post disease onset. The PCR cycle threshold values did not appear to associate with the N antigen concentrations (Figure 1a).

**Figure 1.**
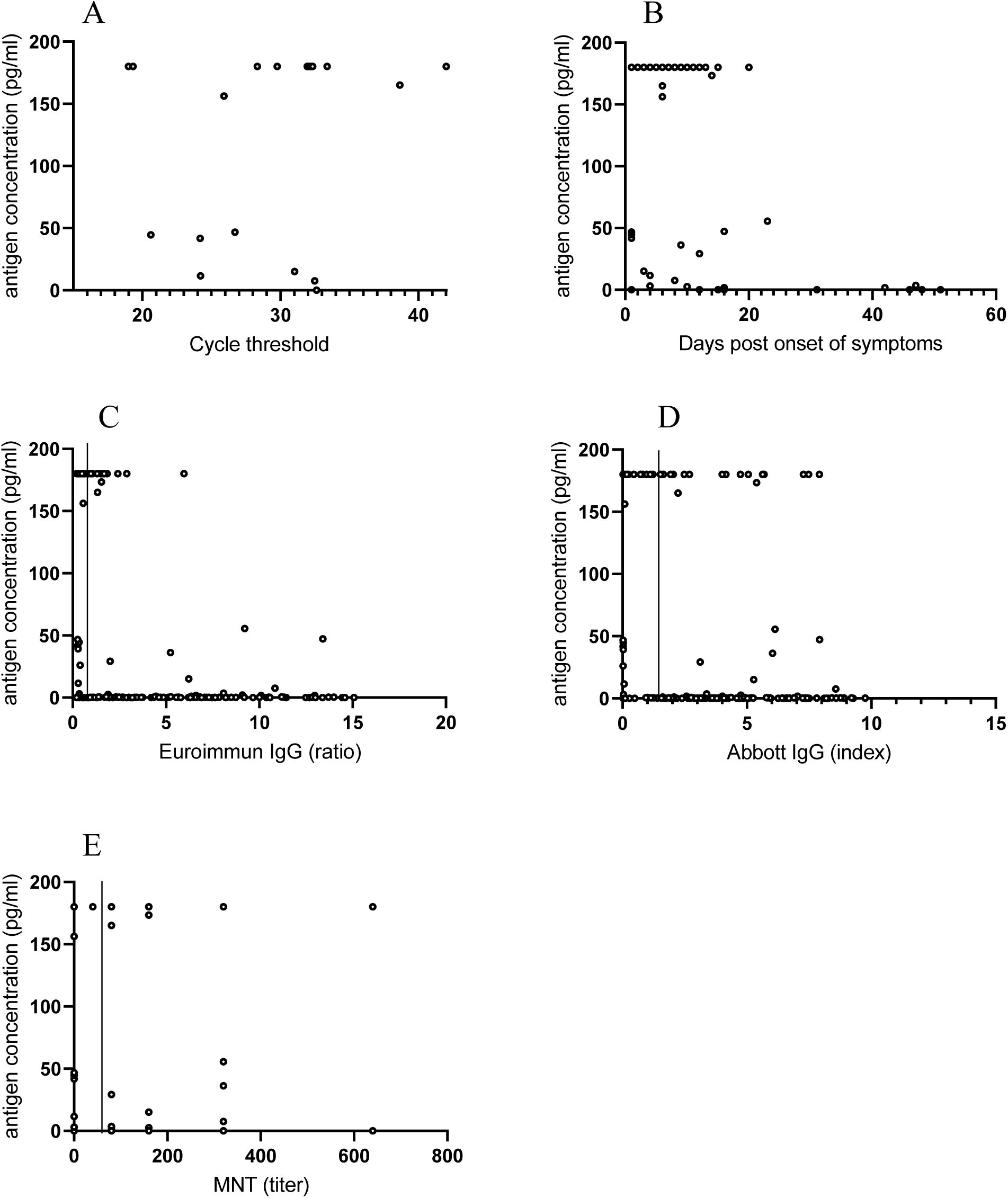
Serum antigen concentrations in relation to the timing of sampling and other tests. Serum antigen concentrations of >180 pg/ml are depicted as 180 pg/ml. A. Serum antigen concentrations compared to the Ct-value of a URT PCR sample obtained on the same day. B. Serum antigen concentrations relative to the timing of sampling after the onset of symptoms. C. Correlation of serum antigen concentration with the Euroimmun IgG assay results. The vertical line indicates the cut-off value of the Euroimmun assay D. Antigen concentrations in comparison with the.Abbott IgG assay results. The vertical line indicates the cut-off value of the Abbott assay E. Serum antigen concentrations relative to the microneutralization test titer. MNT titers of <40 (negative) are depicted as 0 and MNT titers of >2560 as 2560. Figure created using GraphPad Prism 8.0.1 software.

Using COVID-19 panel A, we calculated test sensitivity in relation to disease onset (Table 1). The sensitivity with specimens retrieved at ≤14 days post onset was 94%, and decreased to 50% with specimens retrieved 15-21 days post onset (Table 1). The N antigen concentrations decreased over time from symptom onset (Figure 1b). The furthest time point for a positive N antigen was observed at 47 days post disease onset (3.5 pg/ml) (Figure 1b).

**Table 1.** Sensitivity of serum antigen test in relation to time post symptom onset

| Time post symptom<br>onset (days) | Time post symptom<br>onset (days, mean<br>[SD]) | N | Sensitivity (%<br>[CI 95%]<sup>a</sup>) |
| --- | --- | --- | --- |
| ≤7 | 4.6 (2.3) | 26 | 96.2 (80.4-99.9) |
| 8-14 | 10.4 (1.7) | 24 | 91.7 (73.0-99.0) |
| ≤14 | 7.4 (3.5) | 50 | 94.0 (83.5-98.7) |
| 15-21 | 16.3 (1.9) | 6 | 50.0 (11.8-88.2) |
| ≤21 | 8.4 (4.4) | 56 | 89.3 (78.1-96.0) |
| ≥7 | 16.2 (12.5) | 41 | 75.6 (59.7-87.6) |
| ≥14 | 28.6 (14.9) | 14 | 42.9 (17.7-71.1) |
| ≥21 | 41.1 (10.3) | 7 | 28.6 (3.7-71.0) |

<sup>a</sup>Clopper-Pearson confidence interval

The median days from disease onset for subgroups in panel A were as follows: N antigen positive (≥2.97 pg/ml), MNT antibody negative (<40 titer) (6 days; 25 cases); N antigen positive, MNT antibody positive (10 days; 31 cases); N antigen negative, MNT antibody positive (37 days; 10 cases) (Figure 2).

**Figure 2.**
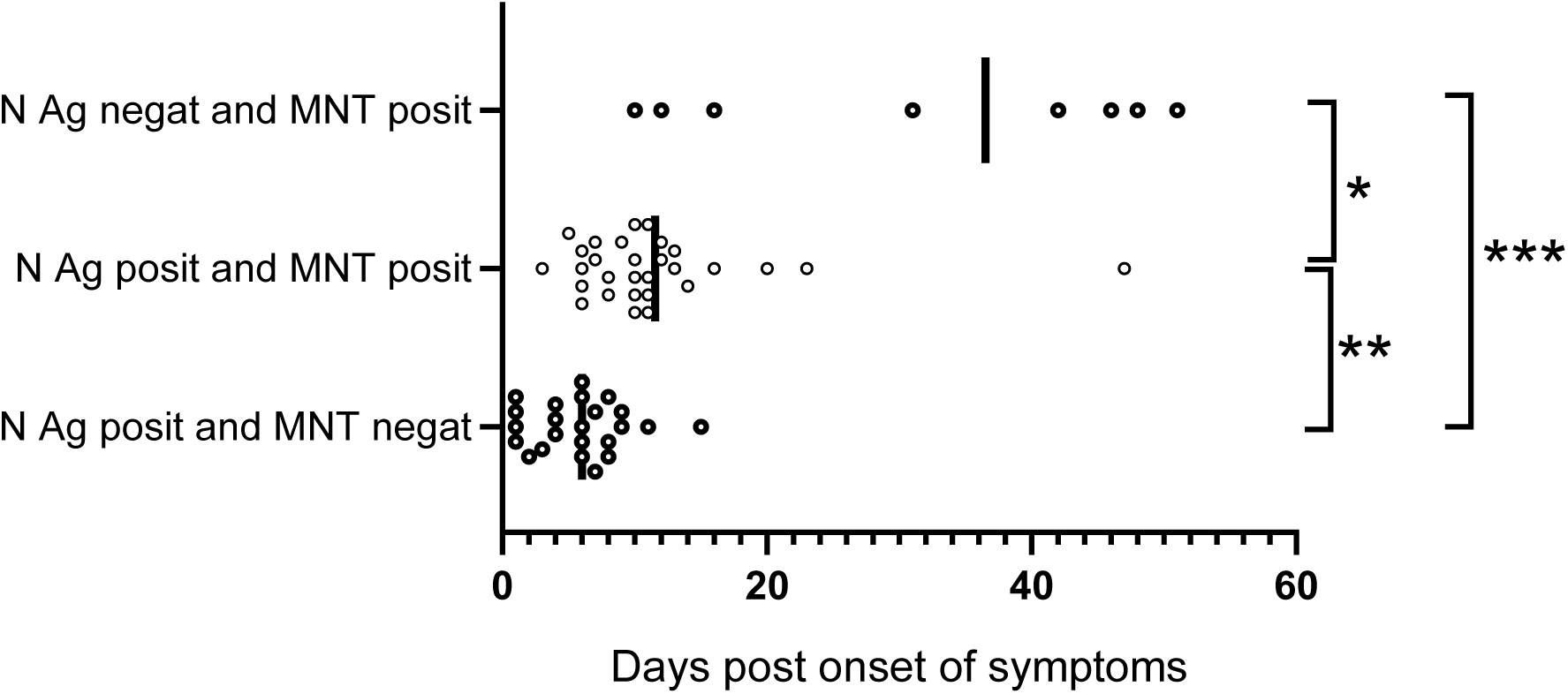
Simultaneous N antigen and MNT test results in relation to symptom onset. N antigen positive and MNT negative, median 6 days. N antigen positive, MNT positive, median 10 days. N antigen negative and MNT positive median. 37 days. Adjusted p-values (Kruskal-Wallis test): *p=0.0317; **p 0.0015, and ***p=<0.0001. Figure created using GraphPad Prism 8.0.1 software.

There appeared to be a decreasing trend in N antigen concentration over increasing Euroimmun anti-S1 IgG test result (Figure 1c), but no such trend was observed over increasing Abbott anti-N IgG test result (Figure 1d), nor the increasing MNT titer (Figure 1e).

Of the COVID-19 panel B, the only N antigen positive serum was retrieved on the date of positive PCR. All other 69 panel B serum specimens were N antigen negative; one was retrieved 5 days before positive PCR in URT, and the others between 12 and 207 days (median 107 days) after positive PCR in URT.

## Discussion

SARS-CoV-2 RNA can be detected for even 4-6 weeks post disease onset in URT samples [9,10]. Subsequently, one of the challenges in contact tracing of PCR positive cases is differentiating between acute COVID-19 disease and convalescent phase. The present study brings forward a potential tool to aid in timing of COVID-19 disease: detection of SARS-CoV-2 N antigen in serum.

Only two previous reports are available on SARS-CoV-2 antigen detection in serum [5,6]. Some antigen tests for the detection of SARS-CoV have been described [11,12]. We showed a very good specificity (98.0%), and a reasonable sensitivity (91.7%; in comparison with simultaneous positive PCR from URT specimens) for the Salocor N antigen test. SARS-CoV-2 N antigenemia appeared to resolve within the first two weeks of illness. Particularly the combination of simultaneous SARS-CoV-2 antigen and antibody testing may provide useful information for timing of disease (Figure 2). These tests can be performed from a single serum specimen in high throughput platforms. While this approach does not replace detection of SARS-CoV-2 RNA from URT, it may provide an additional aid.

Only a limited number of serum specimens was available with information on disease onset, particularly for the convalescent phase. This diminishes the ability of this study to provide a complete timeline for antigenemia in COVID-19. Potential cross-reactivity with seasonal coronaviruses was not assessed in our study.

More data is needed to clarify whether serum N antigen testing could be used to supplement current testing strategies for acute COVID-19. One interesting question is whether there is an association between antigenemia and viral shedding. Besides diagnostic differentiation between acute and convalescent COVID-19, N antigen testing from serum could potentially be deployed in epidemiological screening of asymptomatic infections, e.g. in recently vaccinated populations.

## Data Availability

Researchers may contact study principal investigators to request access to data.

## Acknowledgements

We would like to thank Marita Koivunen (HUS Diagnostic Center, HUSLAB, Helsinki, Finland), Eliisa Kekäläinen (Hus Diagnostic Center and University of Helsinki, Helsinki Finland) and Jesper Kivelä (Hus Diagnostic Center and University of Helsinki, Helsinki Finland).

## Author contributions

MJA: Conceptualization, Investigation, Data curation, Writing – original draft, SKU: Conceptualization, Formal analysis, Resources, Writing – original draft, Writing – review and editing, Supervision, Project administration, SK: Investigation, Writing –review and editing, ML: Resources, Writing – review and editing, Supervision, Project administration HJ: Conceptualization, Formal analysis, Visualization, Resources, Writing – review and editing, Supervision, Project administration, AJJ: Conceptualization, Investigation, Formal analysis, Data curation, Writing – original draft, Supervision, Project administration

