## Supplement 1 for "Detection of SARS-CoV-2 nucleocapsid antigen from serum can aid in timing of COVID-19 infection"

**Salocor SARS-CoV-2 Antigen Quantitative Assay Kit© (Salofa Ltd, Salo, Finland) protocol**

The evaluated Salocor SARS-CoV-2 Antigen Quantitative Assay Kit© is based on a double antibody sandwich method. The kit contains a 96-well plate coated with anti-SARS-CoV-2 N protein antibody. If the serum sample contains SARS-CoV-2 N protein, the N protein attaches both solid phase anti-SARS-CoV-2 N protein antibody and biotin-labelled anti-SARS-CoV-2 N protein antibody added with serum sample. The test procedure is briefly as follows: 50 ul of buffer solution containing biotin-labelled antobody against SARS-CoV-2 N protein was added in each well coated with antibody against SARS-CoV-2 N protein. Next 50 ul of serum sample was added to each well with the exclusion of control well and 5 wells for calibrators included in the kit and added alongside the sample material. After brief mixing, the plate was incubated at 37 °C for 60 minutes followed by washing the wells five times with the washing buffer. 100 ul of horseradish peroxidase-labelled streptavidin was added to each well. After brief mixing, the plate was incubated at 37 °C for 30 minutes followed by the washing step and adding 50 ul of buffer solution containing peroxidase and 50 ul of buffer solution containing 3,3’,5,5’-tetramethylbenzidine. After brief mixing, the plate was incubated at 37 °C for 15 minutes and color reaction was stopped by adding 50 ul of stop solution containing sulfuric acid. Absorbance values were measured at 450 nm with a reference set at 630 nm. The concentration values of samples were then calculated using standard curve with binomial fitting based on different calibrator absorbance values with known SARS-CoV-2 N protein concentrations (pg/ml). Concentration values ≥2,97 pg/ml were interpreted as positive according to the manufacturers´ instructions.
